# Electroconvulsive Therapy with a Memory Reactivation Intervention for Post-Traumatic Stress Disorder: A Randomized Controlled Trial

**DOI:** 10.1101/2020.10.10.20210450

**Authors:** Victor M. Tang, Kathleen Trought, Kristina M. Gicas, Mari Kozak, Sheena A. Josselyn, Zafiris J. Daskalakis, Daniel M. Blumberger, Daphne Voineskos, Yuliya Knyahnytska, Yuan Chung, Young Zhou, Moshe Isserles, Albert H.C. Wong

## Abstract

**Introduction:** Post-traumatic Stress Disorder (PTSD) often does not respond to available treatments. Memories are vulnerable to disruption during reconsolidation, and electroconvulsive therapy (ECT) has amnestic effects. We sought to exploit this phenomenon as a potential treatment for PTSD with a clinical trial of patients with PTSD receiving ECT.

**Methods:** Twenty-eight participants with severe depression with comorbid PTSD referred for ECT treatment were randomly assigned to reactivation of a traumatic or non-traumatic memory using script driven imagery prior to each ECT treatment. Primary outcomes were change in scores on the Modified PTSD Symptom Scale - Self Report (MPSS-SR) and the Clinician-Administered PTSD Scale for DSM-5 (CAPS-5). Assessments were completed by blinded raters. Secondary outcomes included a comparison of the change in heart rate while listening to the script.

**Results:** Twenty-five patients who completed a post-ECT assessment were included in the analysis. No significant group differences were found in the MPSS-SR or CAPS-5 scores from pre-ECT to post-ECT or 3-month follow-ups. However, both groups improved at post-ECT and 3-month follow up. Partial eta squared estimates of effect size showed large effect sizes for all outcomes (η^2^ > 0.13). Changes in heart rate were not significantly different between groups or over time.

**Conclusions:** In this RCT, ECT paired with pre-treatment traumatic memory reactivation was not more effective for treating PTSD symptoms than ECT alone. While our primary hypothesis was not supported, our data provides further support for the efficacy of ECT for improving symptoms of PTSD with comorbid depression.

ClinicalTrials.gov

https://clinicaltrials.gov/ct2/show/NCT04027452

Identifier: NCT04027452

## Introduction

Post-traumatic stress disorder (PTSD) is a debilitating mental health condition that may occur following exposure to a traumatic event or series of events. PTSD is prevalent cross-nationally, with large data surveys from the World Health Organization finding the lifetime prevalence to be 3.9%, of which half reported persistent symptoms (1). First line treatment for PTSD includes trauma-focused psychotherapy, with pharmacological interventions such as selective serotonin reuptake inhibitors considered alternative or adjunctive options (2, 3). Despite treatment, many patients have chronic symptoms, with nonresponse rates as high as 50% (4-6). Stable remission for chronic PTSD is rare, reinforcing the need for more effective treatment options (7).

Core features of PTSD include the presence of intrusive memories related to the traumatic event. Recent advances in the understanding of the neurobiology of memory storage and retrieval has led to the search for novel treatment options that specifically target these mechanisms. Of particular interest in the treatment of PTSD is the reconsolidation hypothesis, which states that long-term memories return to a labile state when reactivated by a memory cue and are subsequently vulnerable to modification by pharmacological, behavioural or psychological interventions (8-10). Many human and animal studies have used propranolol to disrupt the reconsolidation of aversive memories, and both have reported promising results (11). Randomized controlled trials (RCT) in patients with PTSD have found that traumatic memory reactivation coupled with propranolol administration significantly decreases PTSD symptoms and physiologic responses (12, 13). However, memory reconsolidation occurs within a short time window that is difficult to target precisely with oral medications (11).

Electroconvulsive therapy (ECT), in which a generalized tonic-clonic seizure is induced under general anesthesia by direct current stimulation through the scalp, is a highly effective treatment for depression (14). The main side effect and drawback with ECT is amnesia, both anterograde and retrograde, affecting primarily autobiographical details surrounding the course of treatment (15). We sought to harness this undesirable side effect as a potentially therapeutic intervention to disrupt the problematic memories associated with PTSD symptoms.

The impetus for the current study was the findings of Kroes et al. (10). They reported that ECT significantly disrupts the recall of a recently-learned emotionally aversive story if the memory was cued immediately prior to ECT treatment. This memory disruption was specific to the reactivated memory, as control memories that were not reactivated remained intact. Similarly, a case report on a single patient with multiple traumas found that conversation-based reactivation of one traumatic event followed by ECT led to decreased recollection and PTSD symptoms associated with that specific trauma (16). Early preclinical studies also support this concept and demonstrate that electroconvulsive shock is effective in disrupting the reconsolidation of reactivated memories in rodents (17).

To our knowledge there are no published clinical studies to assess the therapeutic effects of using ECT to disrupt the reconsolidation of traumatic memories in patients with PTSD. In this report we describe a single-blind RCT of traumatic memory reactivation followed by ECT for PTSD symptoms compared to reactivation of a neutral, non-traumatic memory. Based on existing neurobiological theories and evidence, we anticipated that ECT administered immediately after the reactivation of a traumatic memory would reduce traumatic memory-related symptoms through disruption of reconsolidation of that memory. Thus, we specifically hypothesized that there would be greater improvement in PTSD symptoms for the traumatic memory exposure group compared to the neutral memory group. This trial represents an important effort to translate clinically relevant neuroscientific knowledge into novel therapeutic options for patients with severe PTSD.

## Methods

This two-group, single-blind RCT was approved by the Research Ethics Board at the Centre for Addiction and Mental Health (CAMH) in Toronto, Canada (REB protocol 063/2015, ClinicalTrials.gov Identifier: NCT04027452). All patients provided written informed consent prior to participating in the study. The trial followed the Consolidated Standards of Reporting Trials (CONSORT) guidelines.

### Sample Recruitment

Patients were recruited from January 2016 until April 2020. During this time period, all inpatients and outpatients with severe depression or suicidality who were referred for ECT treatment at CAMH were pre-screened for eligibility to participate in the study. Patients referred to ECT for depression represent a severe and treatment resistant population that did not respond to standard pharmacological or psychological treatments for depression. Pre-screening was conducted by consulting psychiatrists after approval for ECT. If patients were suitable and interested, they were referred to a research analyst to obtain consent and formally assess eligibility. Patients were required to be aged 18 years or older, deemed suitable for ECT treatment at CAMH and report traumatic memories that cause them distress. Patients were excluded if they were not approved for ECT or if their ECT treatment course had already started prior to study initiation. Other psychiatric diagnoses were not part of the inclusion or exclusion criteria, however they were noted and assessed. Significant treatment effects were found in the previous study by Kroes et al. (10) with 13 participants per group, thus we aimed for a target of at least 13 participants in each arm of the present study.

### Intervention

To re-activate traumatic experiences, we used script-driven imagery (SDI), a method originally described by Pitman et al. (18) and subsequently used in studies with a similar design to the present study (19). In short, each participant was asked to complete a script preparation form describing their traumatic experience, as well as choose five physical symptoms from a given list that accompanied the experience. The same form was also completed for a neutral experience, which we standardized to be a typical morning routine. A research analyst then transformed the subjects’ written descriptions into approximately 125 word long “exposure script” in the second person, present tense. Scripts also incorporated the chosen five physical symptoms. The same study investigator audio recorded all of the scripts in a neutral tone, in order to standardize the scripts in tone and length. Patients were randomized in a 1:1 ratio to traumatic and neutral memory exposure groups using an online randomization tool, which was kept confidential from primary investigators.

Patients attended their ECT sessions as scheduled by their ECT psychiatrist. The clinical indication for ECT and the treatment protocol, including electrode placement and number of sessions, was determined by the clinical care team. Immediately prior to each ECT session, participants listened to the audio scripts through high quality earphones connected to an iPod. Scripts were listened to immediately after IV placement and before anesthesia administration. All recordings were two minutes in length. The first 30 seconds of the recording were instructions, followed by a 30 second silence period to measure physiological baseline. Next, the exposure scripts were played for 30 seconds, where participants were instructed to imagine the described experience. Finally, there was another 30 second silence period where participants were instructed to continue imaging the experience.

ECT was delivered through the MECTA spECTrum 5000Q with a fixed 800 milliamps parameter setting. The stimulus titration method was used to determine the seizure threshold of each individual patient. In the right unilateral setting, stimulus intensity was set to 6 times seizure threshold, to ultrabrief pulse width of 0.3-0.37 milliseconds. Bilateral treatments were given at 1.5 times seizure threshold, and 1.0 millisecond standard pulse width. The anesthetic agent was methohexital 0.75mg-1.0mg/kg IV and the muscle relaxant succinylcholine 0.5-0.75mg/kg IV.

### Psychophysiological Measures

Previous studies have found larger physiologic responses during traumatic imagery in PTSD groups compared to non-PTSD (20). Additionally, psychophysiologic response was found to be significantly smaller in subjects who received post-traumatic memory reactivation with propranolol versus placebo (13). In our study, we assessed the psychophysiologic response by continuously recording participants’ heart rate with an ECG monitor as they listened to the scripts.

### Assessments

At the initial visit, participants underwent a structured evaluation of their psychiatric diagnoses and quantification of their trauma-related symptoms using the Mini International Neuropsychiatric Interview (MINI), Life Events Checklist (LEC), Modified PTSD Symptom Scale Self Report (MPSS-SR) and the Clinician Administered PTSD Scale for DSM-5 (CAPS-5). MPSS-SR is a brief self-report measure that assesses both frequency and severity of PTSD symptoms (21). CAPS-5 is a structured clinical interview to determine diagnosis of PTSD based on DSM-5 (22). Both tools have been found to be reliable and valid in assessing PTSD symptom severity (22, 23). At the post-ECT visit (i.e., the end of the prescribed course of ECT) and 3-month follow-up visit, trauma-related symptoms were captured using MPSS-SR and CAPS-5.

### Statistical Analysis

All analyses were performed using the Statistical Package for the Social Science (SPSS) version 26.0 (SPSS Inc., IBM Corp., Armonk, USA), unless otherwise specified. Continuous variables were examined for normality using visual inspection of histograms and qq-plots. Univariate descriptive analyses were performed by comparing the groups on sociodemographic and clinical characteristics using t-tests or the non-parametric Mann-Whitney U test for continuous variables, and chi-square or Fisher’s exact tests for categorical variables.

### Primary Analyses

Primary outcome measures were changes in MPSS and CAPS-5 total scores from pre- to post-ECT. To determine whether the traumatic memory group showed significantly greater change in symptoms on the primary outcome measures, two separate mixed model ANOVAs (analysis of variance) were conducted with pre- and post-ECT scores entered as the within-subjects factor and memory exposure group entered as the between-subjects factor. Partial eta squared was reported as the estimate of effect size defined by cut-offs of .01 (small), .06 (medium), and .13 (large). The alpha level was set to .025 to adjust for multiple testing. The assumption of homogeneity of regression slopes was examined.

### Secondary Analyses

Using the same procedures described for the primary analyses, mixed model ANOVAs were conducted using pre-ECT, post-ECT, and 3-month follow-up MPSS and CAPS-5 total scores as the outcomes. Additional exploratory analyses were conducted to examine how symptoms changed between groups on the CAPS-5 subscales (intrusion, avoidance, cognition and mood, arousal and reactivity, distress or impairment, global severity, and total PTSD symptoms). Relationships between PTSD symptoms with the LEC score, age of trauma, and length of ECT were also explored (Method S1 in Supplement).

Mean heart rate (HR) was calculated across four contiguous blocks of time that corresponded with the four phases of the memory reactivation paradigm (instructions, baseline, script, and imagery). Mean baseline HR was subtracted from mean imagery HR, and the difference score (HRdiff) was used in subsequent analyses. To examine whether mean HRdiff varied between the traumatic memory and neutral memory groups, and whether HRdiff changed across ECT sessions, a linear mixed effects model was implemented with a random intercept and fixed slope. Only two participants had more than 17 ECT sessions, and therefore these data from later time points were excluded to reduce the risk of bias when missing data is imputed using the full information maximum likelihood approach. The HRdiff score was entered as the repeated measures outcome, whereas group and group*time interactions were entered as the predictors. In follow-up models, MPSS and CAPS-5 change scores were entered as fixed effects predictors to examine whether change in symptoms was associated with a corresponding change in HRdiff across ECT sessions. Analyses were performed in R (R Core Team, 2019) using the *lme4* package (24).

## Results

Flow of participants and reasons for drop out are included in Figure 1. Twenty-eight participants referred for ECT were enrolled and randomized to receive either traumatic or neutral memory reactivation. All participants that completed a post-ECT assessment were included in the analysis: 14 in the traumatic memory group and 11 in the neutral memory group. Three participants in the traumatic memory group discontinued the intervention prematurely and continued to complete the post-ECT assessment; 3 participants in the neutral memory group discontinued the study and did not complete the post-ECT assessment. Twenty one participants (traumatic memory group = 11, neutral memory group = 10) also completed a 3-month follow-up assessment. One participant’s MPSS data at baseline was deemed invalid and another participant did not complete the CAPS-5 at the 3-month follow-up. These cases were excluded from their respective mixed model ANOVAs. Two participants were also missing data for education. Groups were comparable on sociodemographics and clinical characteristics with the exception of a moderately higher number of ECT treatments in the non-traumatic memory group (Table 1).

**Table 1.** Sample Characteristics by Treatment Group.

| Variable | Traumatic Memory Group, (n = 14) | Neutral Memory Group, (n = 11) | Test statistic (p-value) |
| --- | --- | --- | --- |
| Age, mean (SD) | 37.1 (9.2) | 41.7 (12.3) | t = -1.07 (.296) |
| Education, n(%)<sup>a</sup> |  |  |  |
| Less than high school | 1 (7.1) | 0 (0.0) | --- |
| Completed high school | 2 (14.2) | 0 (0.0) | --- |
| Some post-secondary | 5 (35.7) | 5 (45.5) | --- |
| Completed post-secondary | 6 (42.9) | 6 (54.5) | --- |
| Number of ECT treatments, mean (SD) | 10.8 (3.4) | 14.3 (3.1) | t = -2.64 (.015) |
| Number of bilateral ECT treatments, median (IQR) | 2.5 (7) | 0.0 (9) | U = 0.01 (.790) |
| Age at first trauma | 13.6 (8.2) | 15.4 (9.0) | t = -0.50 (.623) |
| LEC total, mean (SD) | 9.0 (2.5) | 8.2 (3.2) | t = 0.72 (.480) |
| MDD Current, n(%) | 13 (92.9) | 11 (100.0) | --- |
| MDD with Psychosis,<br>n(%) | 3 (21.4) | 2 (18.2) | OR = 1.23<br>(1.00) |
| Psychosis, n(%) | 2 (14.3) | 1 (9.1) | OR = 1.67<br>(1.00) |
| Suicide risk, n(%) |  |  |  |
| Low | 1 (7.1) | 0 (0.0) | --- |
| Moderate | 3 (21.4) | 2 (18.2) | --- |
| High | 10 (71.4) | 9 (81.8) | --- |
| PTSD, n(%) | 14 (100.0) | 11 (100.0) | --- |
| Alcohol Dependence, n(%) | 3 (21.4) | 3 (27.3) | OR = 0.73<br>(1.00) |
| Substance Dependence,<br>n(%) | 2 (14.3) | 3 (27.3) | OR = 0.44<br>(.623) |
| GAD, n(%) | 11 (78.6) | 8 (72.7) | OR = 1.38<br>(1.00) |

*U* = Mann-Whitney U test. ECT = electroconvulsive therapy; IQR = interquartile range; LEC = Life Events Checklist; MDD = major depressive disorder; PTSD = post-traumatic stress disorder; GAD = generalized anxiety disorder; OR = odds ratio, computed for Fisher's Exact test. --- = does not meet criteria for 2x2 contingency table.
<sup>a</sup>*n* = 13 for Group X, *n* = 10 for Group Y.

**Figure 1.**
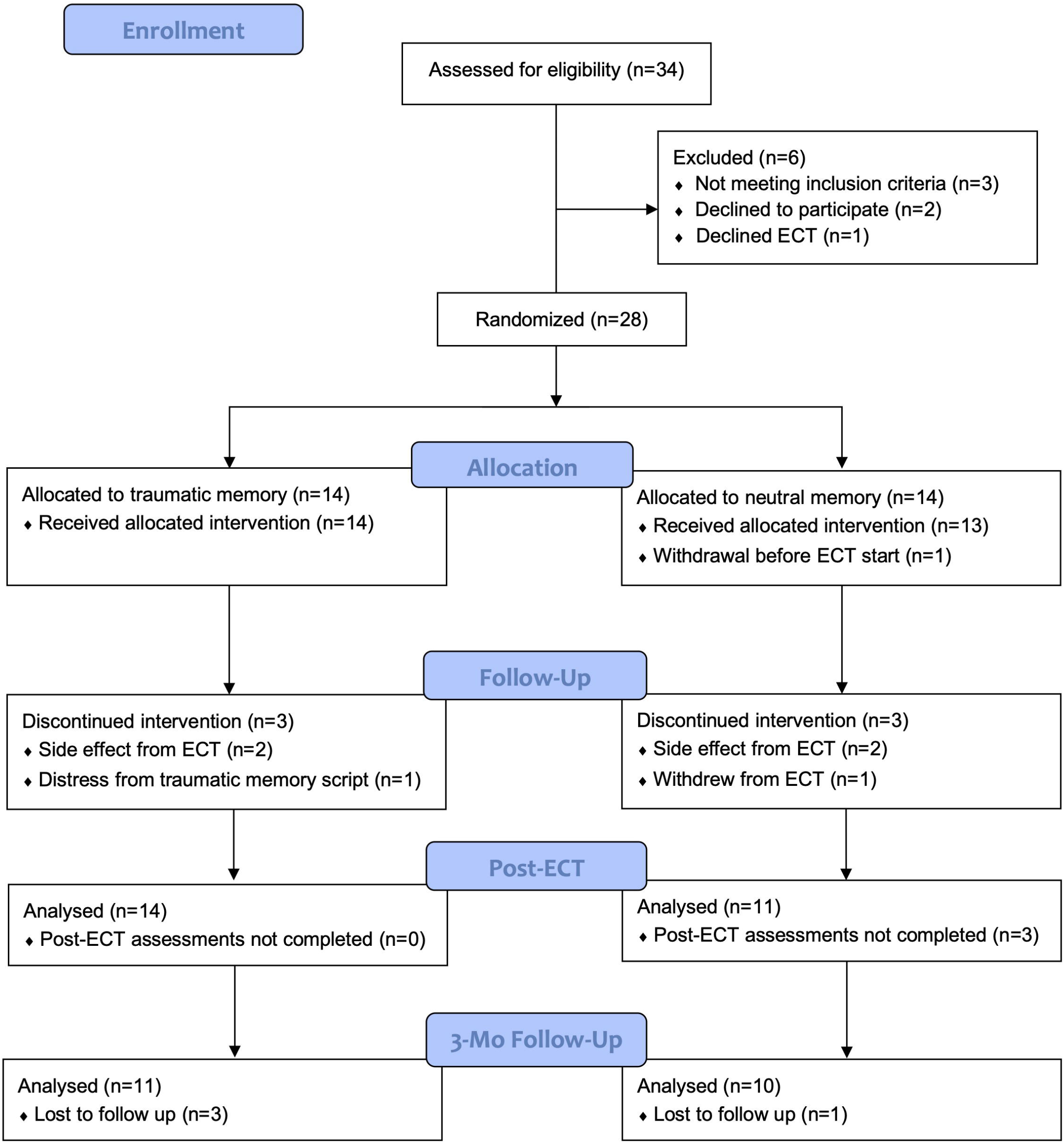
CONSORT flow diagram for participant enrolment and dropouts. CONSORT, Consolidated Standards of Reporting Trials; ECT, Electroconvulsive therapy.

### Primary Analysis

#### PTSD Treatment Outcomes

A main effect was evident in both models, indicating a significant mean decrease in MPSS total scores (F = 30.51, p < .001, η^2^ = .581) and CAPS-5 total scores (F = 22.31, p < .001, η^2^ = .492) from pre-ECT to post-ECT. However, there was no significant group by time interaction for either MPSS-SR (F = 0.59, p = .449, η^2^ = .026) or CAPS-5 scores (F = 0.28, p = .604, η^2^ = .012). Mean MPSS-SR and CAPS-5 scores at each assessment are summarized by group in Figures 2 and 3, respectively.

**Figure 2.**
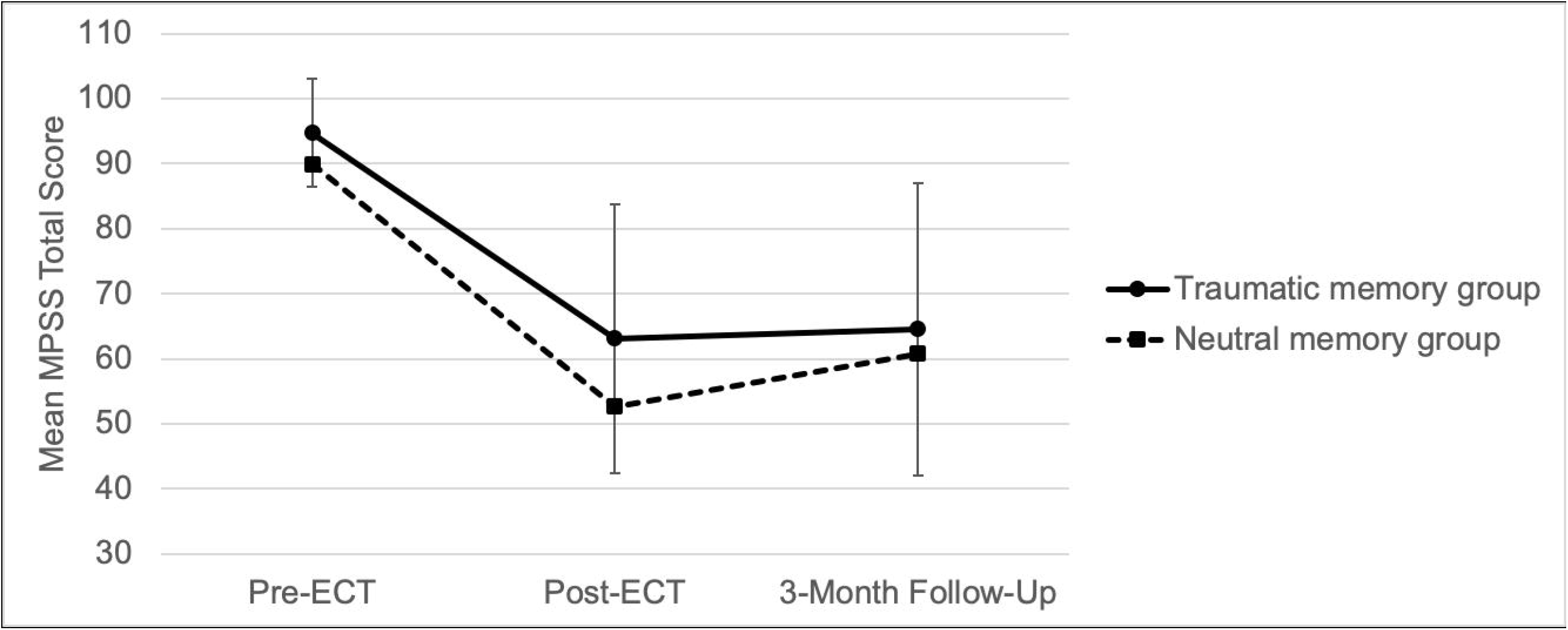
Modified PTSD Symptom Scale-Self Report (MPSS-SR) mean scores presented for Pre-ECT, Post-ECT, and 3-Month Follow-Up time points. PTSD, Post-Traumatic Stress Disorder; ECT, Electroconvulsive Therapy. Error bars represent 95% confidence intervals.

**Figure 3.**
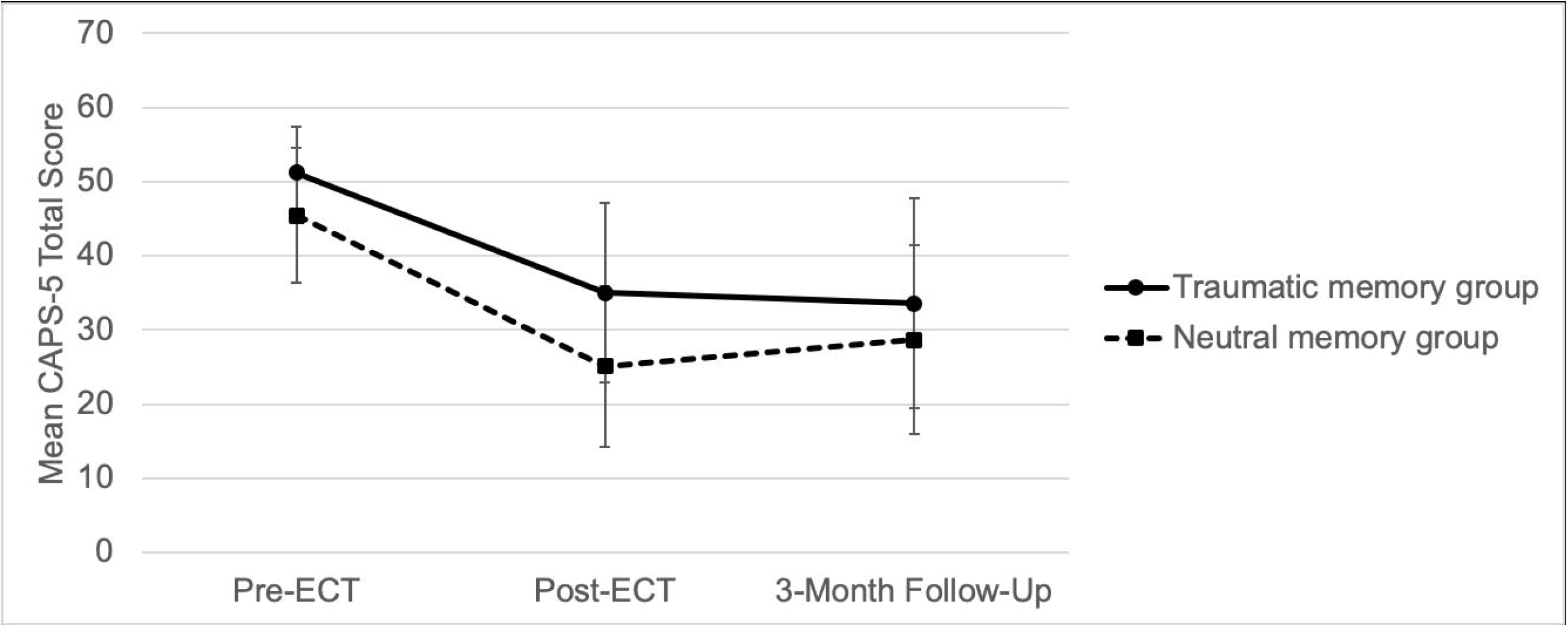
Clinician Administered PTSD Scale for DSM-5 (CAPS-5) mean scores presented for Pre-ECT, Post-ECT, and 3-Month Follow-Up time points. PTSD, Post-Traumatic Stress Disorder; ECT, Electroconvulsive Therapy. Error bars represent 95% confidence intervals.

### Secondary Analysis

#### PTSD Outcomes at 3-Month Follow Up

Results of the mixed model ANOVAs using symptom data from all three assessment points were similar to the previous results whereby there was evidence of significant overall change in MPSS scores (F = 23.23, p < .001, η^2^ = .563) and in CAPS-5 scores (F = 17.50, p < .001, η^2^ = .493), but mean symptom change did not differ by group (MPSS - Group*Time: F = .05, p = .956, η^2^ = .002; CAPS-5 - Group*Time: F = .08, p = .928, η^2^ = .004).

#### PTSD Symptom Subscale Outcomes

Exploratory analyses of PTSD symptom subscales (intrusion, avoidance, cognition and mood, and arousal and reactivity symptoms), or for items reporting clinically significant distress or impairment in social, occupational, or other important areas of functioning and global severity on the CAPS-5 showed evidence of significant overall change in all domains across all timepoints, but no significant group differences were found (Table S1).

#### Correlations with Characteristics of Trauma History

In the combined sample, no significant correlations were observed between LEC total score and change in MPSS total scores (ρ = .144, p = .502) or change in CAPS-5 total scores (ρ = .235, .257). There was also no difference between age at first trauma groups in MPSS change scores (<= 12 years: 32.5(27.0); >12 years: 32.5 (32.7); t = -.04, p = .997) or CAPS-5 change scores (<= 12 years: 11.5 (15.1); >12 years: 21.6 (19.4); t = −1.42, p = .170). Examination of correlations between symptom change scores at 3-month follow-ups revealed a medium-to-large, positive association between the LEC total score and CAPS-5 change score in the combined sample (ρ = .461, p = .041). No association was observed for LEC total score and MPSS change score (ρ = .184, p = .438), nor was there any difference in change scores between age at first trauma groups for MPSS scores (<= 12 years: 33.0 (20.5); >12 years: 26.6 (23.3); t = .64, p = .530) or CAPS-5 scores ((<= 12 years: 16.4 (9.0); >12 years: 17.8 (18.0); t = -.22, p = .829). In addition, number of ECT sessions showed a non-significant medium-to-large negative association with change in CAPS-5 scores at 3 months (ρ = -.433, p = .056, but not with change in CAPS-5 scores post-ECT (ρ = -.255, p = .280). No associations were observed between the number of ECT sessions and change in MPSS scores post-ECT (ρ = -.102, p = .635) or at 3 months (ρ = -.210, p = .374). All correlations, including those with the trauma and neutral groups presented separately, are available in Table S2.

#### Psychophysiological Outcomes

There was a non-significant difference with a large effect size between the traumatic and neutral memory group during the imagery portion of the script (Figure S1). In the mixed effects model examining HRdiff response (e.g., HR imagery - HR baseline), there was no difference between the two groups (b = -.173, SE = .941, p = .854) at the pre-ECT assessment. There was also no evidence of change in HR response over time (b = .111, SE = .074, p = .134) and this did not differ by group (b = -.068, SE = .096, p = .483). In follow-up models, change in MPSS scores (b = -.001, SE = .002, p = .703) and CAPS-5 scores (b = .003, SE = .003, p = .386) were not associated with change in HR.

## Discussion

In this study, we examined the efficacy of traumatic memory reactivation prior to ECT treatments in reducing symptoms of PTSD by interfering with reconsolidation. Although ECT is known to cause retrograde amnesia for a short period surrounding treatment, no statistically reliable effect of our memory reactivation procedure on PTSD symptoms as measured by the CAPS-5 and MPSS-SR was found. However, both groups showed a reduction of PTSD symptoms for both the MPSS and CAPS-5 total scores, with treatment effects sustained after 3-months. The change in scores corresponded with large effects sizes that represent clinically important differences (25). Supplementary analyses suggested that improvement in PTSD symptoms may be associated with a greater number of different trauma exposures over the lifetime and fewer ECT sessions. While these correlations should be interpreted with some caution due to the small sample size, these preliminary findings highlight the potential important role that trauma history characteristics may play in treatment response. Heart rate patterns did not differ between the trauma and neutral memory groups nor did they significantly change over the course of ECT. Our findings suggest that ECT is effective in the treatment of PTSD but without any added benefit of a traumatic memory reactivation intervention.

A number of important factors could have influenced our results, including the nature of individual trauma, effectiveness of our memory reactivation procedure, and the ECT parameters. In the Kroes et al. (10) study that successfully disrupted experimentally-created memories with reactivation before ECT, those memories were discrete and artificial. In our sample, most had early life trauma consisting of multiple different types of events, some of which may not be easily defined as discrete events (see LEC in Table 1). These multiple, overlapping and prolonged traumatic events are difficult to selectively reactivate in a single script. This is corroborated by the case report of a patient with PTSD from numerous traumatic events who received a course of ECT with memory reactivation of just one of those events prior to each treatment (16). PTSD symptoms related to the reactivated memory subsided and there was modest improvement of overall CAPS-5 scores. However, there was still a significant burden of PTSD symptoms remaining, interpreted as the ongoing effects of the multiple other traumas.

Older fear memories are more stable and less plastic (26). The average traumatic memory reactivated in our study was over 20 years old (Table 1), much older than the experimentally-created memories in previous studies (10). Older memories may require more intensive reconsolidation disruption (27) and our dose of ECT may have been insufficient. Furthermore, increases in the stress response to the memory and the “training strength” (i.e., the number of times a conditioning stimulus is paired with an unconditioned stimulus) is associated with greater resistance to manipulation (28, 29). In a treatment-resistant PTSD sample such as this, traumatic memories are associated with severe levels of distress, and those with repeated, enduring forms of traumatic exposures likely represent greater “training strength”, as would be modeled in animal studies.

Lastly, it is possible that our script driven imagery protocol was inadequate and required optimization. Measurement of autonomic arousal suggested engagement with the script, however this was not significant (Figure S1). Many studies have shown that when a memory is reactivated there needs to be some degree of updated content to be learned during the retrieval (i.e., a prediction error) in order for reconsolidation to occur (30). Another possibility is that longer memory re-exposures are more likely to induce reconsolidation (28). In a positive trial of propranolol for blocking reconsolidation of traumatic memories, the trauma reactivation protocol took approximately 10-20 minutes (12), substantially longer than our exposure script. Thus, future reactivation protocols could involve modifications of the memory (e.g. a cognitive reappraisal of the memory) or a longer exposure time.

The current study reinforces the challenges related to applying neuroscientific principles of memory reconsolidation for real-world, clinical populations. The majority of published studies on reconsolidation of aversive memories are in healthy human subjects as opposed to clinical samples (11). Relatedly, Wood et al. (31) reported on 3 studies in patients with PTSD using pharmacological agents to impair reconsolidation, all of which found no significant effects on clinical or psychophysiological outcomes. Our study recruited patients with complex courses of illness, to which previous data on healthy subjects may not apply (32).

An important finding of our study is that ECT produced a significant reduction in CAPS-5 and MPSS-SR scores in both groups, with clinically significant effect sizes. Due to ethical considerations, this study was not designed to recruit patients being referred to ECT for the intention of treating their PTSD given that there currently does not exist an adequate evidence base to support ECT for PTSD. Data from this study provides support for the use of ECT in PTSD with comorbid MDD. There is only one other prospective clinical trial of ECT for PTSD, an open label trial of 20 patients (33) which found a 70% response rate. Our study provides an additional advance by having a larger sample size of 25 patients, as well as follow-up assessments at 3 months. There have been retrospective studies that suggest ECT is equally effective in those with depression and comorbid PTSD compared with depression only (34), and may be superior to pharmacotherapy only (35). Further research on ECT for PTSD is warranted.

There are other limitations to be acknowledged. In this study the number of ECT treatments, stimulus parameters, electrode placement, were not standardized for the treatment of PTSD or the reactivation protocol. Future studies will benefit from determining whether dose or type of ECT treatment is more likely to be effective when delivered in with memory reactivation. For example, bilateral ECT may prove to be more effective at disrupting reconsolidation as it is known to cause greater memory side effects than unilateral ECT (36). There was also variability in the psychiatric comorbidity and other concurrent psychiatric treatments received. However, we believe that less stringent enrollment criteria allowed us to test the reconsolidation hypothesis in a realistic clinical population for which these novel treatments would eventually be intended. This could be improved in the future by including symptom scales for other symptom domains such as for MDD. Lastly, future studies should further optimize patient characteristics, such as focusing on recruiting patients with adult single-incident traumas which may address potentially confounding developmental effects (37).

The strength of the present study includes the novel study design targeting the neurobiological processes underlying traumatic memories central to PTSD. There is a growing literature on human laboratory and clinical studies on disrupting aversive memory reconsolidation using pharmacological or behavioural interventions, with the goal of translating these principles into effective treatment options for clinical populations. We present the first clinical trial evaluating the effectiveness of this approach using ECT in PTSD. Our participants represented a population of high clinical interest, as they had not responded to standard pharmacological or psychological treatments, and development of novel treatments are greatly needed.

In conclusion, our results did not show any significant clinical benefits of a traumatic memory reactivation protocol to target the reconsolidation process, but ECT in general may be an effective treatment for symptoms of PTSD. Future research may continue to elucidate the optimal parameters for which ECT and memory reactivation can be used to disrupt traumatic memories and ultimately lead to more clinically significant effects.

## Data Availability

Data available on request due to privacy/ethical restrictions.

## Acknowledgments

We would like to thank Dr. Yuan Chung, Young Zhou, Tania Jain, Amelia Srajer, Xing Gao, Amy Freeman and Justin Ryk for their assistance in data collection.

## References

1. Koenen K, Ratanatharathorn A, Ng L, McLaughlin K, Bromet E, Stein D, et al.Posttraumatic stress disorder in the world mental health surveys. Psychological medicine. 2017;47(13):2260–74.

2. Watkins LE, Sprang KR, Rothbaum BO. Treating PTSD: A review of evidence-based psychotherapy interventions. Frontiers in Behavioral Neuroscience. 2018;12:258.

3. Jonas DE, Cusack K, Forneris CA, Wilkins TM, Sonis J, Middleton JC, et al.Psychological and pharmacological treatments for adults with posttraumatic stress disorder (PTSD). 2013.

4. Murdoch M, Sayer NA, Spoont MR, Rosenheck R, Noorbaloochi S, Griffin JM, et al. Long-term outcomes of disability benefits in US veterans with posttraumatic stress disorder. Archives of General Psychiatry. 2011;68(10):1072–80.

5. Steenkamp MM, Litz BT, Hoge CW, Marmar CR. Psychotherapy for military-related PTSD: A review of randomized clinical trials. Jama. 2015;314(5):489–500.

6. Schottenbauer MA, Glass CR, Arnkoff DB, Tendick V, Gray SH. Nonresponse and dropout rates in outcome studies on PTSD: Review and methodological considerations. Psychiatry: Interpersonal and Biological Processes. 2008;71(2):134–68.

7. Shalev AY. Posttraumatic stress disorder and stress-related disorders. Psychiatric Clinics. 2009;32(3):687–704.

8. Nader K, Schafe GE, Le Doux JE. Fear memories require protein synthesis in the amygdala for reconsolidation after retrieval. Nature. 2000;406(6797):722–6.

9. Nadel L, Land C. Memory traces revisited. Nature reviews neuroscience. 2000;1(3):209–12.

10. Kroes MC, Tendolkar I, Van Wingen GA, Van Waarde JA, Strange BA, Fernández G. An electroconvulsive therapy procedure impairs reconsolidation of episodic memories in humans. Nature neuroscience. 2014;17(2):204–6.

11. Bolsoni LM, Zuardi AW. Pharmacological interventions during the process of reconsolidation of aversive memories: A systematic review. Neurobiology of Stress. 2019;11:100194.

12. Brunet A, Saumier D, Liu A, Streiner DL, Tremblay J, Pitman RK. Reduction of PTSD symptoms with pre-reactivation propranolol therapy: a randomized controlled trial. American Journal of Psychiatry. 2018;175(5):427–33.

13. Brunet A, Orr SP, Tremblay J, Robertson K, Nader K, Pitman RK. Effect of post-retrieval propranolol on psychophysiologic responding during subsequent script-driven traumatic imagery in post-traumatic stress disorder. Journal of psychiatric research. 2008;42(6):503–6.

14. The U. Efficacy and safety of electroconvulsive therapy in depressive disorders: a systematic review and meta-analysis. The Lancet. 2003;361(9360):799–808.

15. Fink M, Taylor MA. Electroconvulsive therapy: evidence and challenges. Jama. 2007;298(3):330–2.

16. Gahr M, Schönfeldt-Lecuona C, Spitzer M, Graf H. Electroconvulsive therapy and posttraumatic stress disorder: first experience with conversation-based reactivation of traumatic memory contents and subsequent ECT-mediated impairment of reconsolidation. The Journal of neuropsychiatry and clinical neurosciences. 2014;26(3):E38–E9.

17. Misanin JR, Miller RR, Lewis DJ. Retrograde amnesia produced by electroconvulsive shock after reactivation of a consolidated memory trace. Science. 1968;160(3827):554–5.

18. Pitman RK, Orr SP, Forgue DF, de Jong JB, Claiborn JM. Psychophysiologic assessment of posttraumatic stress disorder imagery in Vietnam combat veterans. Archives of General Psychiatry. 1987;44(11):970–5.

19. Isserles M, Shalev AY, Roth Y, Peri T, Kutz I, Zlotnick E, et al. Effectiveness of deep transcranial magnetic stimulation combined with a brief exposure procedure in post-traumatic stress disorder–a pilot study. Brain stimulation. 2013;6(3):377–83.

20. Orr SP, Metzger LJ, Pitman RK. Psychophysiology of post-traumatic stress disorder. Psychiatric Clinics. 2002;25(2):271–93.

21. Falsetti SA, Resnick HS, Resick PA, Kilpatrick DG. The Modified PTSD Symptom Scale: A brief self-report measure of posttraumatic stress disorder. The Behavior Therapist. 1993.

22. Weathers FW, Bovin MJ, Lee DJ, Sloan DM, Schnurr PP, Kaloupek DG, et al. The Clinician-Administered PTSD Scale for DSM–5 (CAPS-5): Development and initial psychometric evaluation in military veterans. Psychological assessment. 2018;30(3):383.

23. Ruglass LM, Papini S, Trub L, Hien DA. Psychometric properties of the modified posttraumatic stress disorder symptom scale among women with posttraumatic stress disorder and substance use disorders receiving outpatient group treatments. Journal of Traumatic Stress Disorders & Treatment. 2014;4(1).

24. Bates D, Mächler M, Bolker B, Walker S. Fitting Linear Mixed-Effects Models using lme4. 2015; 67 (1): 48. Epub 2015-10-07. https://doi.org/10.18637/jss.v067.i01; 2015.

25. Stefanovics EA, Rosenheck RA, Jones KM, Huang G, Krystal JH. Minimal clinically important differences (MCID) in assessing outcomes of post-traumatic stress disorder. Psychiatric Quarterly. 2018;89(1):141–55.

26. Elsey JW, Kindt M. Breaking boundaries: optimizing reconsolidation-based interventions for strong and old memories. Learning & Memory. 2017;24(9):472–9.

27. Bustos SG, Maldonado H, Molina VA. Disruptive effect of midazolam on fear memory reconsolidation: decisive influence of reactivation time span and memory age. Neuropsychopharmacology. 2009;34(2):446–57.

28. Suzuki A, Josselyn SA, Frankland PW, Masushige S, Silva AJ, Kida S. Memory reconsolidation and extinction have distinct temporal and biochemical signatures. Journal of Neuroscience. 2004;24(20):4787–95.

29. Wang S-H, de Oliveira Alvares L, Nader K. Cellular and systems mechanisms of memory strength as a constraint on auditory fear reconsolidation. Nature neuroscience. 2009;12(7):905–12.

30. Sevenster D, Beckers T, Kindt M. Retrieval per se is not sufficient to trigger reconsolidation of human fear memory. Neurobiology of learning and memory. 2012;97(3):338–45.

31. Wood NE, Rosasco ML, Suris AM, Spring JD, Marin M-F, Lasko NB, et al. Pharmacological blockade of memory reconsolidation in posttraumatic stress disorder: three negative psychophysiological studies. Psychiatry research. 2015;225(1-2):31–9.

32. Soeter M, Kindt M. High trait anxiety: A challenge for disrupting fear memory reconsolidation. PloS one. 2013;8(11):e75239.

33. Margoob MA, Ali Z, Andrade C. Efficacy of ECT in chronic, severe, antidepressant-and CBT-refractory PTSD: an open, prospective study. Brain stimulation. 2010;3(1):28–35.

34. Kaster TS, Goldbloom DS, Daskalakis ZJ, Mulsant BH, Blumberger DM. Electroconvulsive therapy for depression with comorbid borderline personality disorder or post-traumatic stress disorder: A matched retrospective cohort study. Brain stimulation. 2018;11(1):204–12.

35. Ahmadi N, Moss L, Simon E, Nemeroff CB, Atre-Vaidya N. EFFICACY AND LONG-TERM CLINICAL OUTCOME OF COMORBID POSTTRAUMATIC STRESS DISORDER AND MAJOR DEPRESSIVE DISORDER AFTER ELECTROCONVULSIVE THERAPY. Depression and anxiety. 2016;33(7):640–7.

36. Sackeim HA, Prudic J, Devanand DP, Nobler MS, Lisanby SH, Peyser S, et al. A prospective, randomized, double-blind comparison of bilateral and right unilateral electroconvulsive therapy at different stimulus intensities. Archives of General Psychiatry. 2000;57(5):425–34.

37. Klengel T, Mehta D, Anacker C, Rex-Haffner M, Pruessner JC, Pariante CM, et al. Allele-specific FKBP5 DNA demethylation mediates gene–childhood trauma interactions. Nature neuroscience. 2013;16(1):33–41.

