## Supplementary material for "Electroconvulsive Therapy with a Memory Reactivation Intervention for Post-Traumatic Stress Disorder: A Randomized Controlled Trial": Method S1

### Supplemental Information

#### Method S1 in Supplement

Spearman's rho correlations and t-tests were used to examine whether MPSS and CAPS-5 symptom change from pre-ECT to 3-month follow-up was associated with the LEC total score and age at first trauma. Associations between total number of ECT sessions and symptom change were also explored. Given the small sample size and increased risk of a Type II error, adjustments for multiple testing were not applied for the secondary analyses. In follow-up, mean MPSS and CAPS-5 total change scores were computed by subtracting pre-ECT from post-ECT scores. An index of lifetime exposure to trauma was calculated by computing the total number of exposures from a possible 17 different events measured by the LEC. To examine whether aspects of trauma history were associated with responsiveness to ECT treatment, Spearman's rho correlations were used to examine associations between the derived LEC total score and symptom change scores in the combined sample (traumatic and neutral memory groups). Due to a skewed distribution, age at first trauma was dichotomized as less than or equal to 12 years or greater than 12 years. T-tests were used to compare the age at first trauma groups on symptom change scores.
