## Supplementary material for "Electroconvulsive Therapy with a Memory Reactivation Intervention for Post-Traumatic Stress Disorder: A Randomized Controlled Trial": Figure S1

**Supplemental Figure**

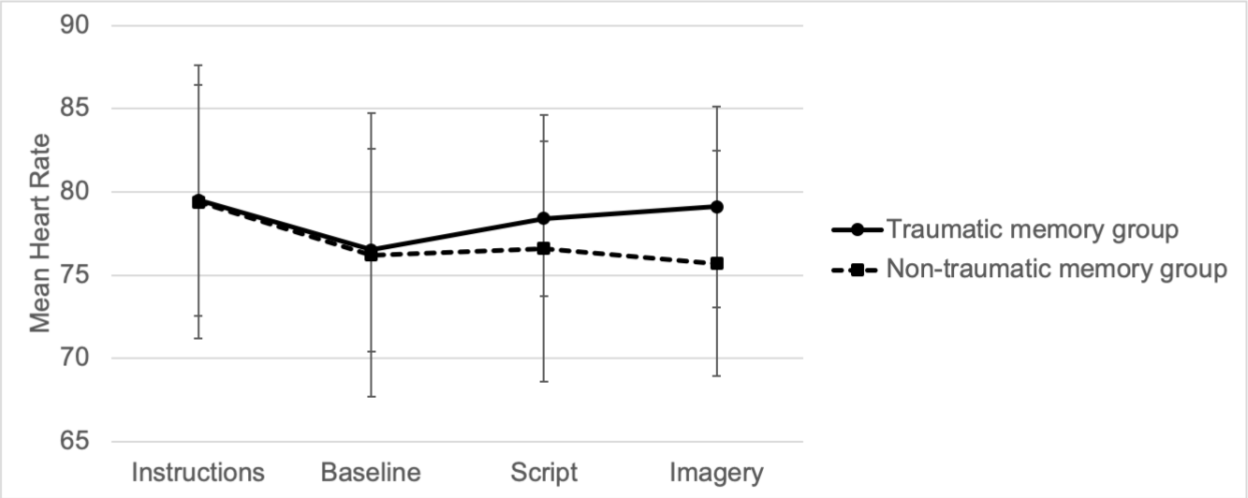


Figure S1. Mean heart rate across sections of the script driven imagery for ECT treatment at baseline to validate the degree of emotional arousal and thus engagement with the script. The second ECT treatment was selected due to the potential stress associated with attending the first treatment of ECT, and subsequent sessions were not included to exclude potential effects of habituation over time. Error bars represent 95% confidence intervals.
