## Supplementary material for "Electroconvulsive Therapy with a Memory Reactivation Intervention for Post-Traumatic Stress Disorder: A Randomized Controlled Trial": Table S1

**Supplemental Table**

Table S1. Mean change in Clinician-Administered PTSD Scale for DSM-5 (CAPS-5) subscale scores by group.

| CAPS-5 Subscale<br>Group | Pre-ECT Mean(SD) | Post-ECT Mean(SD) | 3-month Mean(SD) | F-statistic (p-value) | Effect size |
|---|---|---|---|---|---|
| PTSD symptom number* | | | | | |
| Traumatic | 15.6 (2.7) | 11.9 (6.6) | 10.6 (6.1) | Time: 22.38 (<.001) | .493 |
| Non-traumatic | 15.4 (2.6) | 8.4 (4.9) | 9.9 (5.4) | Group*Time: 2.10 (.160) | .084 |
| Intrusion | | | | | |
| Traumatic | 14.1 (3.4) | 9.0 (6.5) | 8.6 (7.4) | Time: 16.60 (<.001) | .419 |
| Non-traumatic | 10.8 (4.5) | 5.5 (4.1) | 6.4 (4.4) | Group*Time: 0.01 (.910) | .001 |
| Avoidance | | | | | |
| Traumatic | 5.7 (2.6) | 4.1 (2.9) | 3.9 (3.1) | Time: 8.38 (.008) | .267 |
| Non-traumatic | 5.4 (1.9) | 3.1 (1.7) | 7.4 (5.4) | Group*Time: 0.28 (.602) | .012 |
| Cognition and mood | | | | | |
| Traumatic | 20.1 (5.1) | 12.4 (7.8) | 12.7 (7.6) | Time:37.22 (<.001) | .618 |
| Non-traumatic | 17.7 (5.6) | 8.8 (6.6) | 11.7 (7.8) | Group*Time: 0.19 (.665) | .008 |
| Arousal and reactivity | | | | | |
| Traumatic | 11.4 (3.0) | 9.5 (5.6) | 8.5 (5.4) | Time: 6.83 (.016) | .229 |
| Non-traumatic | 11.5 (4.5) | 7.8 (6.2) | 7.4 (5.4) | Group*Time: 0.72 (.406) | .030 |
| Distress or impairment** | | | | | |
| Traumatic | 9.9 (1.6) | 6.3 (3.9) | 7.2 (2.8) | Time: 36.25 (<.001) | .612 |
| Non-traumatic | 9.6 (1.4) | 4.6 (2.5) | 6.0 (2.6) | Group*Time: 1.00 (.329) | .042 |
| Global severity | | | | | |
| Traumatic | 3.1 (1.0) | 2.0 (1.5) | 2.1 (1.0) | Time: 14.10 (.001) | .380 |
| Non-traumatic | 3.1 (0.7) | 2.0 (0.9) | 2.2 (1.0) | Group*Time: 0.01 (.946) | .000 |

*The total number of items on the CAPS-5 that are scored ≥2 indicating clinically significant severity.
**The sum of CAPS-5 items G23 (overall subjective distress), G24 (impairment in social functioning), and G25 (impairment in occupational or other important area of functioning)

Table S2. Correlations between PTSD symptom change, trauma, and number of ECT sessions

| Correlations Between PTSD Change and Number of Trauma Types (LEC) |
|---|
| LEC total number of exposures and MPSS-SR change post-ECT<br>Trauma (n = 13): Spearman's rho = .076, p = .805<br>Neutral (n = 11): Spearman's rho = .265, p =.431<br>Fisher's z-test: z = -.41, p = .681<br>Full sample: Spearman's rho = .144, p =.502<br><br>LEC total number of exposures and CAPS-5 total change post-ECT<br>Trauma (n = 14): Spearman's rho = .143, p = .625<br>Neutral (n = 11): Spearman's rho = .235, p = .479<br>Fisher's z-test: z = -.29, p = .771<br>Full sample: Spearman's rho = .235, p = .257<br><br>LEC total number of exposures and MPSS-SR change at 3 months<br>Trauma (n = 10): Spearman's rho = .508, p = .134<br>Neutral (n = 10): Spearman's rho = .080, p = .827<br>Fisher's z-test: z = .90, p = .369<br>Full sample: Spearman's rho = .184, p =.438<br><br>LEC total number of exposures and CAPS-5 change at 3 months<br>Trauma (n = 11): Spearman's rho = .431, p = .186<br>Neutral (n = 9): Spearman's rho = .408, p = .276<br>Fisher's z-test: z = .05, p = .959<br>Full sample: Spearman's rho = .461, p = .041 |

Correlations Between PTSD Change and Age of Index Trauma

Age of trauma (<=12 yo vs. >12 yo) and MPSS-SR change post-ECT
Trauma (n = 13): Point biserial r = .082, p = .789
Neutral (n = 10):
Point biserial r = -.454, p = .188
Fisher's z-test: z = 1.16, p = .256
Full sample: Spearman's rho = -.066, p =.766

Age of trauma (<=12 yo vs. >12 yo) and CAPS-5 total change post-ECT
Trauma (n = 14): Point biserial r = .443, p = .112
Neutral (n = 10):
Point biserial r = .244, p = .496
Fisher's z-test: z = .47, p = .639
Full sample: Spearman's rho = .337, p = .107

Age of trauma (<=12 yo vs. >12 yo) and MPSS-SR change at 3 months
Trauma (n = 10): Point biserial r = -.213, p = .554
Neutral (n = 9): Point biserial r = .000, p = 1.00
Fisher's z-test: z = -.39, p = .697
Full sample: Spearman's rho = -.096, p =.695

Age of trauma (<=12 yo vs. >12 yo) and CAPS-5 total change at 3 months
Trauma (n = 11): Point biserial r = .000, p = 1.00
Neutral (n = 9): Point biserial r = .348, p = .359
Fisher's z-test: z = -.67, p = .501
Full sample: Spearman's rho = .122, p = .609

Correlations Between PTSD Change and Number of Total ECT Treatments

Total number of ECT treatments and MPSS-SR change post-ECT

Trauma (n = 13): Spearman's rho = -.172,  p = .575
Neutral (n = 11): Spearman's rho = -.186,  p = .584
Fisher's z-test: z = .03, p = .976
Full sample: Spearman's rho = -.102, p =.635

Total number of ECT treatments and CAPS-5 total change post-ECT

Trauma (n = 14): Spearman's rho = -.403,  p = .153
Neutral (n = 11): Spearman's rho = -.117,  p = .732
Fisher's z-test: z = -.67, p = .505
Full sample: Spearman's = .280

Total number of ECT treatments and MPSS-SR change at 3 months

Trauma (n = 10): Spearman's rho = -.477,  p = .163
Neutral (n = 10): Spearman's rho = -.095,  p = .794
Fisher's z-test: z = -.79, p = .428
Full sample: Spearman's rho = -.210, p =.374

Total number of ECT treatments and CAPS-5 total change at 3 months

Trauma (n = 11): Spearman's rho = -.638,  p = .035
Neutral (n = 9): Spearman's rho = -.308,  p = .420
Fisher's z-test: z = -1.01, p = .312
Full sample: Spearman's rho = -.433, p = .056
